# Resolving inter- and intra-patient heterogeneity in *NPM1-*mutated AML at single-cell resolution

**DOI:** 10.1101/2024.12.10.24317471

**Authors:** E Onur Karakaslar, Eva M Argiro, Nadine E Struckman, Ramin HZ Shirali, Jeppe F Severens, M Willy Honders, Susan L Kloet, Hendrik Veelken, Marcel JT Reinders, Marieke Griffioen, Erik B van den Akker

**Author notes:** **Correspondence** Erik B van den Akker Marieke Griffioen Leiden University Medical Center Albinusdreef 2, Leiden, Netherlands 2333 ZA.

## Abstract

*NPM1-*mutated AML is one of the largest entities in international classification systems of myeloid neoplasms, which are based on integrating morphologic and clinical data with genomic data. Previous research, however, indicates that bulk transcriptomics-based subtyping may improve prognostication and therapy guidance. Here, we characterized the heterogeneity in *NPM1*-mutated AML by performing single-cell RNA-sequencing and spectral flow cytometry on 16 AML belonging to three distinct subtypes previously identified by bulk transcriptomics. Using single-cell expression profiling we generated a comprehensive atlas of *NPM1*-mutated AML, collectively reconstituting complete myelopoiesis. The three *NPM1*-mutated transcriptional subtypes showed consistent differences in the proportions of myeloid cell clusters with distinct patterns in lineage commitment and maturational arrest. In all samples, malignant cells were detected across different myeloid cell clusters, indicating that *NPM1-*mutated AML are heavily skewed but not fully arrested in myelopoiesis. Same-sample multi-color spectral flow cytometry recapitulated these skewing patterns, indicating that the three *NPM1*-mutated subtypes can be consistently identified across platforms. Moreover, our analyses highlighted differences in the abundance of rare hematopoietic stem cells suggesting that skewing occurs early in myelopoiesis. To conclude, by harnessing single-cell RNA-sequencing and spectral flow cytometry, we provide a detailed description of three distinct and reproducible patterns in lineage skewing in *NPM1*-mutated AML that may have potential relevance for prognosis and treatment of patients with *NPM1*-mutated AML.

**KEY MESSAGES:**

- *NPM1*-mutated AML shows strong intra- and interpatient heterogeneity with malignant cells skewed rather than fully arrested at different maturation stages in myelopoiesis.
- Single-cell RNA sequencing and spectral flow cytometry revealed recurrent patterns in proportions of malignant myeloid cells with distinct patterns in lineage commitment and maturational arrest.

**GRAPHICAL ABSTRACT:** 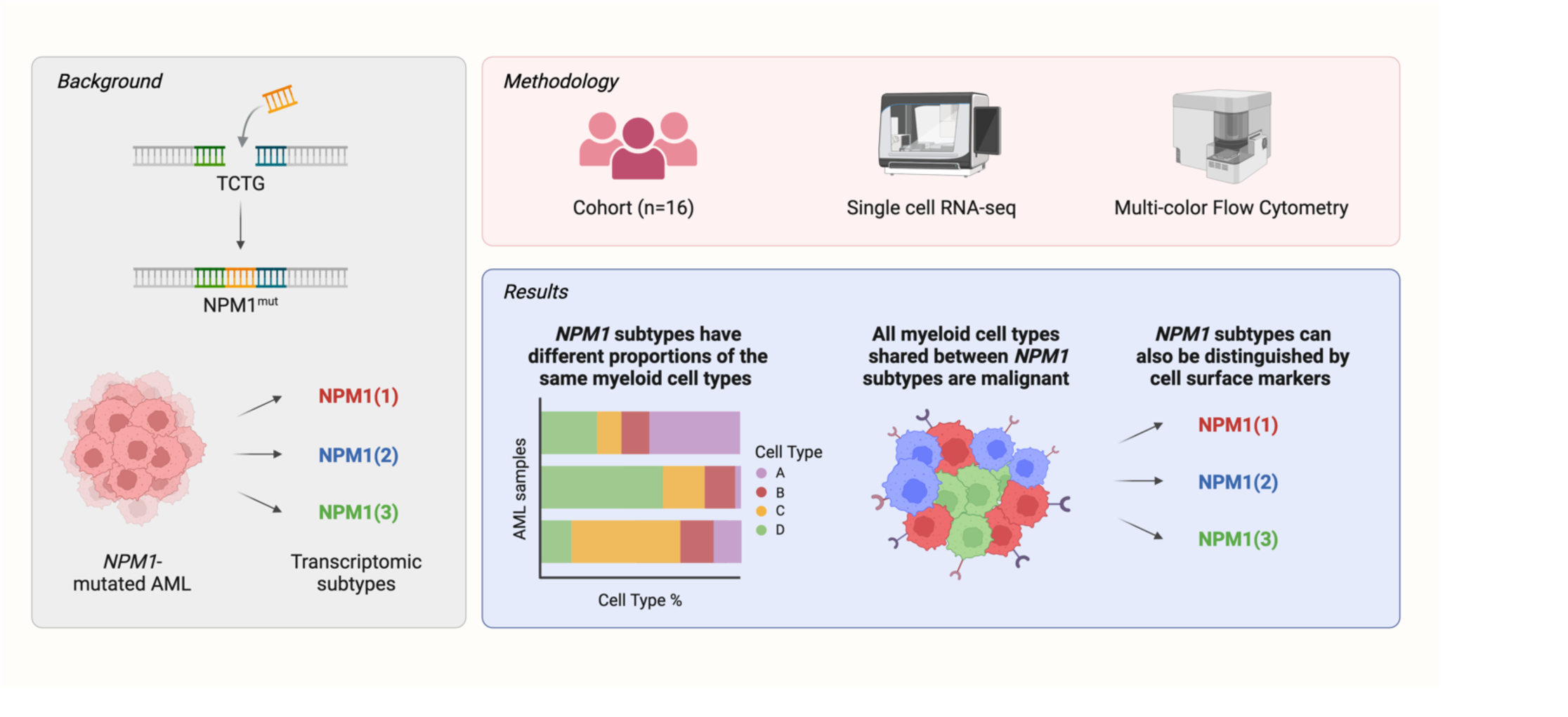

## INTRODUCTION

Acute Myeloid Leukemia (AML) is a heterogeneous malignant disease characterized by uncontrolled proliferation of immature myeloid precursor cells in the bone marrow. The latest WHO and ICC classifications stratify adult AML into various categories that are mainly based on distinct recurrent genetic aberrations, with one of the largest entities being *NPM1-*mutated AML^1,2^. A 4 base pair frameshift mutation in *NPM1* is sufficient to drive malignant transformation. Although *NPM1*-mutated AML generally have a relatively favorable prognosis^3^, other co-mutations are also important and may affect the prognosis and treatment response of AML patients. For example, patients who acquire *FLT3-ITD* and *DNMT3A*^R882^ co-mutations in addition to the *NPM1* mutation have a significantly worse prognosis^4^.

In line with others who also reported on different gene signatures, we previously identified three distinct subtypes of *NPM1*-mutated AML by bulk-transcriptomics, each showing distinct ex-vivo drug sensitivity profiles, and enrichments for different co-mutations^5–8^. Patients with the transcriptional NPM1(1) subtype had mutually exclusive co-mutations with either *IDH1/2* or *TET2*, whereas AML samples with NPM1(2) or NPM1(3) subtypes showed strong enrichments for *FLT3-ITD* or *DNMT3A*^R882^ co-mutations, respectively. These observations raise the question whether the three distinct transcriptional subtypes in *NPM1*-mutated AML are the result of different co-mutational patterns or whether the three subtypes reflect progenitor cells arrested at different stages in myelopoiesis with variable susceptibility to acquire certain type of co-mutations.

Most large-scale gene expression studies, including ours, leveraged RNA sequencing data in bulk of AML samples in which healthy and malignant hematopoiesis may co-exist^9,10^. Recent single-cell RNA sequencing revealed that leukemic cells with different maturational arrests carry different gene mutations^10,11^, and that these leukemic cell subsets in AML typically exhibit phenotypes reminiscent of healthy hematopoietic cell types, albeit often accompanied with aberrant characteristics^10,12–14^. Consequently, to advance our understanding of *NPM1*-mutated AML, single-cell analyses are essential for investigating and comparing intra-patient heterogeneity at a resolution that can highlight transcriptional subtypes identified in large-scale bulk sequencing studies.

In this study, we employed single-cell RNA sequencing and spectral flow cytometry to investigate and compare the cellular heterogeneity between 16 *NPM1*-mutated AML selected from the three distinct subtypes previously identified by bulk transcriptomics. Using both single-cell modalities, we identified various leukemic cell clusters resembling different maturation stages during normal hematopoiesis. These leukemic cell clusters were present in different proportions in all three transcriptional subtypes. Leukemic cells resembling early hematopoietic stem and multipotent progenitor cells dominated in two *NPM1* subtypes, whereas leukemic cell subsets with more mature myeloid phenotypes were enriched in the third subtype. Our data support a model in which the three *NPM1* transcriptional subtypes originate from different progenitor cells with variable capacity to produce more differentiated myeloid offspring. Since AML cells with different maturation phenotypes have shown variable *ex-vivo* drug responses, our data may be relevant for the prognosis and treatment of patients and provide preliminary insight into the etiology of *NPM1*-mutated AML.

## RESULTS

### Single-cell atlas of *NPM1*-mutated AML reconstitutes myelopoiesis

To investigate the identity and cellular composition of the three *NPM1* transcriptional subtypes, we performed single-cell RNA sequencing on samples from 16 *NPM1*-mutated AML patients (NPM1(1) n=7; NPM1(2) n=5; NPM1(3) n=4) (Figure 1a). Notably, all patient-derived samples featured the hotspot frameshift mutation in *NPM1*, and co-mutation patterns varied across subtypes as previously described^5^ (Figure 1b, Supplementary Table 1, see Methods).

**Figure 1.**
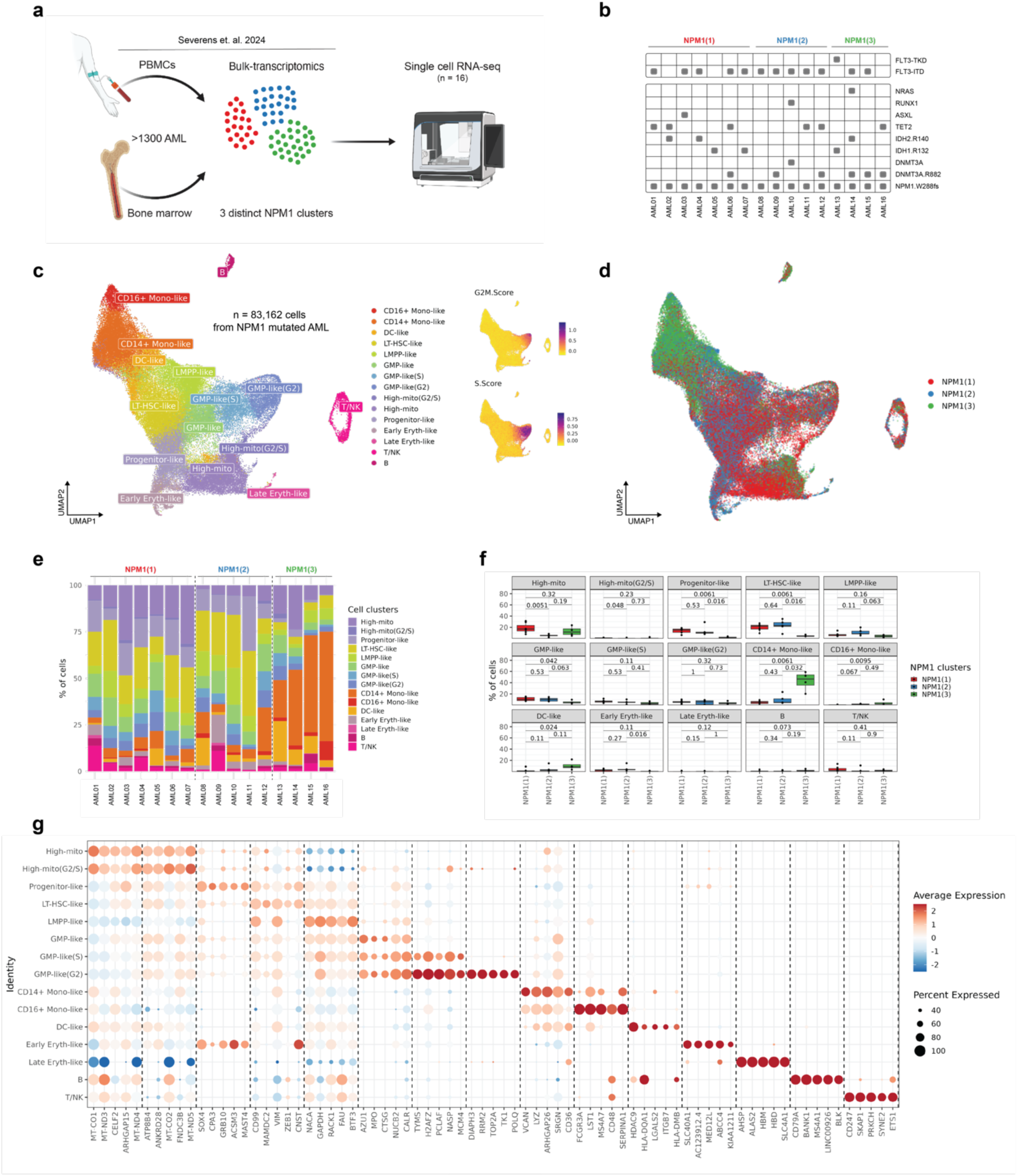
AML heterogeneity between NPM1 subtypes by single-cell RNA-sequencing. **a** Experimental design – 16 AML cases representing the three distinct transcriptional NPM1 subtypes identified by bulk transcriptomics were selected for single cell RNA-sequencing. **b** Gene mutations detected in our cohort of 16 AML cases. **c** UMAP plot showing cells from 16 AML cases (n=83,162) colored by annotated cell types accompanied by cell cycle information **d** UMAP plot colored by NPM1 subtype **e** Barplot showing cell cluster distribution per patient. Patients were grouped based on their NPM1 subtypes. **f** Boxplots showing percentages of cell clusters per sample grouped by NPM1 subtypes. Boxplots colored by NPM1 subtype. p values are calculated using the Wilcoxon rank-sum test. **g** Heatmap showing the top 5 up-regulated DEGs per cell cluster. Dot color indicates average expression per cluster and size shows the percentage of cells expressing the respective gene for that cluster.

After stringent quality control and doublet removal, single-cell RNA sequencing (scRNA-seq) data from 83,162 total cells were integrated and clustered into 15 cell clusters (Figure 1c, S1, S2a-d, see Methods). Subsequent annotation of these clusters was performed using various strategies. First, we used Azimuth^15^ to project a healthy bone marrow atlas onto our data (Figure S2e). This allowed us to identify the most resembling cell subset in healthy hematopoiesis for each of our identified clusters. Given that particularly the myeloid lineage is affected by disease, we refer to ‘-like’ subsets for clusters mostly resembling granulocyte-monocyte progenitors (GMPs), monocytes, dendritic cells (DCs), and erythroid cells. While clusters with mature myeloid cell populations, e.g. monocytes and erythrocytes, strongly resembled their healthy counterparts, most clusters with characteristics of progenitor cell populations could not be faithfully projected onto the healthy bone marrow atlas (Figure S2f). We next compared the obtained cell clusters to a previously published dataset by van Galen *et al.*^12^ containing 16 AML with diverse genetic lesions, including 5 *NPM1*-mutated AML (Figure S2g). Many of our cell clusters resembled one of the 6 cell types previously described in this dataset, including hematopoietic stem cell (HSC)-like, progenitor-like, GMP-like, promonocyte-like, monocyte-like, and conventional dendritic cell (cDC)-like malignant cells.

The combined analysis resulted in 11 myeloid cell clusters including one long-term HSC-like (LT-HSC-like) cluster expressing *CD34*, one lymphoid-primed multipotent progenitor-like (LMPP-like) cluster, one progenitor-like cluster, three GMP-like clusters in different cell cycle states (non-cycling, S, and G2), two monocyte-like clusters (CD14^+^ and CD16^+^ monocytes), one DC-like cluster and two erythrocyte-like clusters (early and late erythrocytes) (Figure S2g, h). In addition, two lymphoid cell subsets clustered separately from the myeloid cells (B-cells and NK/T cells), which is in line with previously published AML studies^5,6^.

Next, we performed additional analyses to explore the identity of two remaining cell clusters with a relatively high mitochondrial RNA content that passed our stringent quality control (Figure S2c). A similar cell cluster was observed in a recent single-cell transcriptomics study including 10 *NPM1*-mutated AML^13^. Since a relatively high content of mitochondrial RNAs may indicate poor data quality, we evaluated to what extent our clustering was affected by variability in mitochondrial content. After adjusting for mitochondrial content, the two high-mito clusters persisted as independent clusters (Figure S2i, j). Moreover, projecting the cells of each individual patient into a separate map revealed that high-mito cells were present within each AML sample (Figure S3). Collectively, these data suggest that the two cell clusters with a high content of mitochondrial RNAs are not the result of poor-quality data of one or a few samples but represent a consistent signal across all AML samples.

Finally, for each cell cluster, differentially expressed genes (DEGs) were identified by comparing its gene expression profile with the other cell clusters (Figure 1g, Supplementary Table 2). We plotted the DEGs identified for high-mito cells using an independent healthy bone marrow atlas^16^ and demonstrated that these cells exhibited signatures reminiscent of early progenitors (Figure S2k). Overall, expression of the DEGs was stronger in the more mature myeloid cell clusters than in the hematopoietic stem cell or progenitor cell clusters. Of note, the DEGs identified for the progenitor-like cell cluster were also expressed by early erythrocyte-like cells, suggesting a common lineage origin and early commitment of progenitor-like cells towards the erythrocyte lineage.

To conclude, our single-cell transcriptomics atlas revealed 13 myeloid cell clusters in *NPM1*-mutated AML representing different maturation signatures and states varying from early to late myeloid differentiation.

### Transcriptional subtypes of *NPM1*-mutated AML exhibit distinct cell type compositions

To investigate whether the cell clusters were exclusively present in one of the three transcriptional *NPM1*-mutated AML subtypes, or whether these cells were present in all three subtypes in different proportions, we checked the distribution of the three *NPM1*-mutated AML subtypes over the UMAP plot (Figure 1d). This analysis showed a clear difference in distribution between the three subtypes due to differences in abundance of specific cell clusters. This observation was further corroborated when the composition of each AML sample was quantified, with samples from the same subtype showing relatively similar cell cluster compositions, and samples from different subtypes exhibiting notable differences in cell cluster composition (Figure 1e, S2l). Moreover, in alignment with our previous observations, NPM1(1) and NPM1(2) samples had more abundant cell clusters with early progenitor phenotypes, whereas cell clusters resembling more mature monocytes and DCs were dominating in NPM1(3) samples (Figure 1f). Of note, although both NPM1(1) and NPM1(2) subtypes were more abundant in cell clusters with early progenitor phenotypes, there were also differences between the two subtypes. For instance, the two high-mito clusters were more abundant in NPM1(1) samples (8-33%, Wilcoxon rank-sum test *P* = 0.005 and *P* = 0.05 for high-mito and -G2, respectively), whereas NPM1(2) samples had higher fractions of LT-HSC-like and LMPP-like cell clusters albeit not significant. Overall, the single-cell transcriptomics data revealed that all 13 cell clusters, representing different myeloid differentiation stages, were present in different proportions in the three *NPM1*-mutated AML subtypes.

### NPM1(2) samples resemble pluripotent hematopoietic stem cells

To further explore the difference in composition between NPM1(1) and NPM1(2) subtypes, we compared the expression of *CD34*, which has been used as marker to annotate LT-HSC-like cells. The data showed that *CD34* expression was mostly restricted to NPM1(2) samples (Figure 2a). To validate this finding, *CD34* expression was analyzed in primary *NPM1*-mutated AML samples from BEAT-AML, an independent bulk-transcriptomics cohort^17^. This analysis confirmed that NPM1(2) samples had higher expression of *CD34* (Wilcoxon rank-sum *P* = 0.0009, Figure 2b, Figure S4a). Since previous studies associated *FLT3-ITD* status with high *CD34* expression^12,18^, we also separately analyzed *NPM1*-mutated AML samples of BEAT-AML based on their *FLT3-ITD* status (Figure S4b). The data showed that *CD34* expression was higher in NPM1(2) samples compared to other subtypes, irrespective of *FLT3-ITD* status (*P* = 0.09).

**Figure 2.**
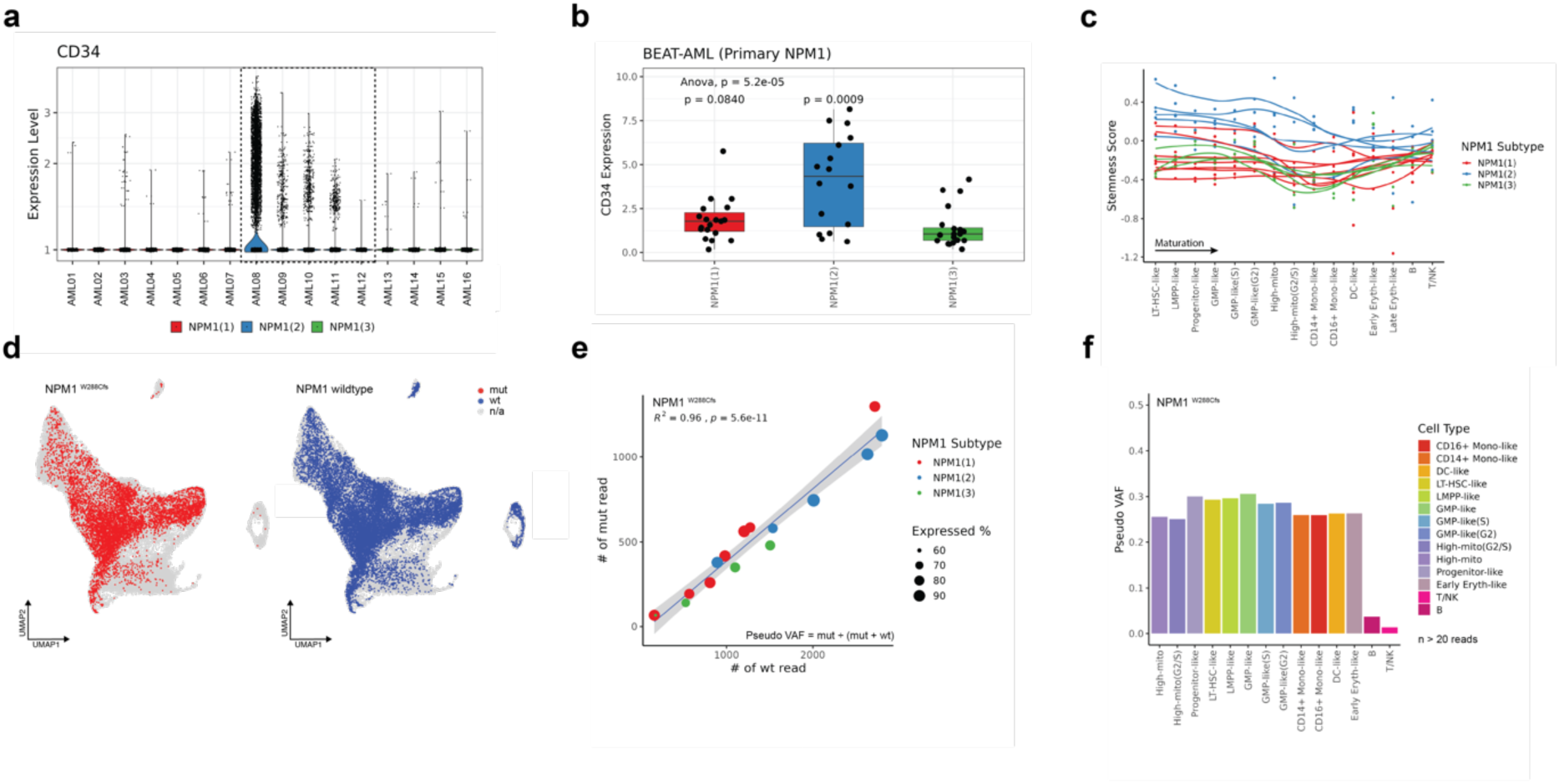
AML in NPM1(2) subtype resemble hematopoietic stem cells or early progenitor cells. **a** Violin plot for *CD34* gene expression per sample. NPM1(2) samples are highlighted. **b** CD34 gene expression of NPM1-mutated AML at diagnosis of the BEAT-AML cohort. p values are calculated using the Wilcoxon rank-sum test with NPM1(3) samples as reference. **c** Stemness score of each cell cluster calculated per sample. To visualize per sample trends smoothed lines were fitted across cell types. Cell clusters are ordered based on their maturation stage. **d** UMAPs plots annotated with cells expressing mutated NPM1^W288Cfs^ (left) or wild type NPM1 allele (right). **e** Scatter plot of mutant (mut) and wildtype (wt) NPM1 allele counts per sample. Samples are colored by their respective NPM1 subtype. **f** Barplot of NPM1^W288Cfs^ pVAF for all cell types, except for late erythrocytes in which less than 20 reads were detected. All myeloid cell clusters are malignant, whereas lymphoid cell clusters mostly lack the mutation.

To further characterize the differences between the *NPM1* clusters, we calculated the stemness score^19^ for our AML samples per cell cluster by creating pseudo-bulks from single-cell data. While the stemness score was shown to decline along the myeloid maturation trajectory, NPM1(2) samples had consistently higher stemness scores across all cell clusters compared to NPM1(1) and NPM1(3) subtypes (Figure 2c). Cross-checking the individual genes contributing to the stemness score revealed that *CD34* expression strongly correlated with stemness score (Figure S4c-d, Pearson *R* = 0.62).

Altogether, our data indicate that NPM1(2) samples have the highest stemness scores and that they seem to retain transcriptional characteristics of pluripotent HSCs along differentiation towards more committed progenitor and mature myeloid cells.

### All myeloid cell clusters in *NPM1*-mutated AML samples are malignant

To investigate whether the identified cell clusters represent malignant cells, we performed variant calling in single-cell RNA-seq data for the known hotspot mutations *NPM1*^WS288Cfs^ and *DNMT3A*^R882H^. Mutated and wildtype *NPM1* reads were detected in all AML samples (Figure 2d, S4e). In the two lymphoid cell clusters (B and T/NK cell clusters), most single-cells had only wildtype *NPM1* transcripts, whereas mutant *NPM1* transcripts were detected in many single-cells in all myeloid cell clusters (Figure S4f). The recall rates of transcripts across cell clusters were concordant with the expression of the *NPM1* gene (Figure S4g).

Read distributions showed that most cells had only one read covering either the mutant or wildtype NPM1 allele (Figure S4h). We therefore determined pseudo variant allele frequencies (pVAF) by calculating the ratio of mutant reads to the total number reads for a given number of cells. All 16 *NPM1*-mutated AML samples (Figure 2e, R^2^ = 0.96, *P* = 5.6x10^-11^) and all myeloid cell clusters (Figure 2f) had similar pVAF. Cell clusters with more mature myeloid phenotypes had slightly lower pVAF compared to early progenitor cell clusters, but these differences were not significant (Figure S4i, *P* = 0.75). This slight decrease in pVAF was also observed when samples were compared between the three NPM1 subtypes, with NPM1(3) samples having overall lower pVAF (Figure S4j). Also, NPM1(3) samples in BEAT-AML had lower VAF_DNA_ than NPM1(1) and NPM1(2) subtypes, but the difference was not statistically significant (Figure S4k). Despite the low number of single cells with *DNMT3A*^R882H^, mutant reads for this hotspot mutation were detected in all cell clusters present in AML samples with this mutation (Figure 1b, Figure S4l). Overall, for each sample, we detected mutant *NPM1* transcripts in all myeloid cell clusters, thereby confirming the malignant origin of the single cells in these clusters.

### Heterogeneity of *NPM1*-mutated AML captured by spectral flow cytometry

To investigate whether *NPM1*-mutated AML from the three transcriptional subtypes also displayed different cell surface markers, we performed spectral flow cytometry using an antibody panel against 19 cell surface markers (Figure 3a, S5a, see Methods). For this purpose, antibodies against well-established hematopoietic stem or progenitor cell and myeloid differentiation markers were used (CD33, CD45, CD34, CD38, CD117, CD45RA, CLL-1, GPR56, CD14, CD16, CD61, CD11c, CD163, CD68, CD85k-LILRB4) as well as antibodies against HLA class I and II molecules (HLA-DR, -DQ, -DP), for which low gene expression was observed particularly in the NPM1(1) subtype^5^ (Figure S5b).

**Figure 3.**
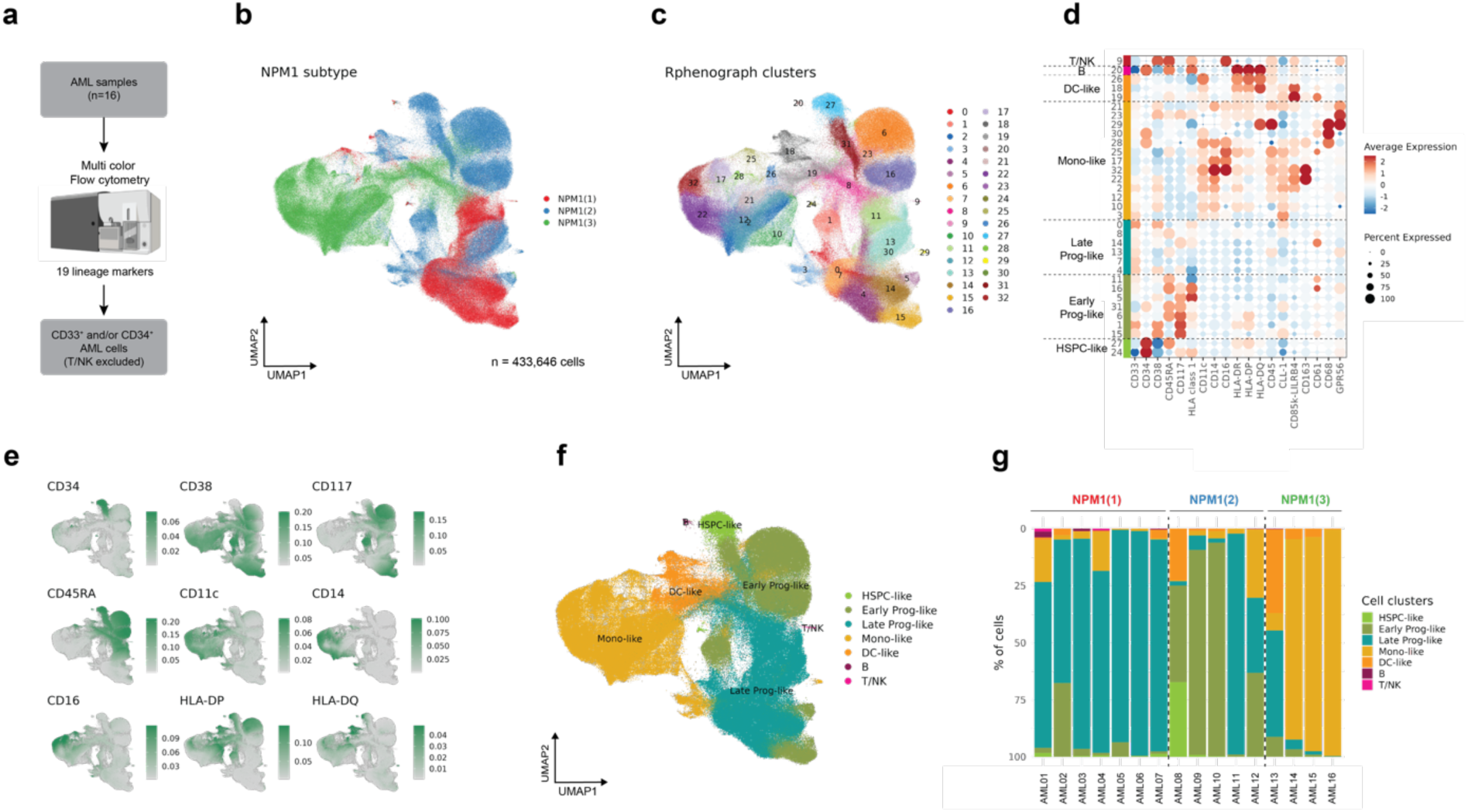
AML heterogeneity between NPM1 subtypes by spectral flow cytometry. **a** Chart illustrating the strategy for spectral flow cytometry. The same 16 AML samples analyzed by single-cell RNA-sequencing were also measured by spectral flow cytometry. **b** UMAP plot of AML cells (n=433,646) measured by spectral flow cytometry data colored by NPM1 subtypes. **c** UMAP plot for 33 *PhenoGraph* clusters (k = 60). **d** Dot plot showing all 19 markers. Each marker is scaled across clusters. Colored bars on the left indicate cell type annotations. **e** Expression of 9 cell surface markers (CD34, CD38, CD117, CD45RA, CD11c, CD14, CD16, HLA-DP and HLA-DQ). **f** UMAP plot colored with cell type annotations. **g** Barplot showing cell type distribution per patient. Patients are grouped based on their NPM1 subtypes.

We used this spectral flow cytometry antibody panel to stain the same set of 16 *NPM1*-mutated AML cases as analyzed by single-cell transcriptomics. After removal of doublets, dead cells and lymphocytes, CD33^+^ and/or CD34^+^ cells were gated, and data were analyzed for a total of 433,646 cells. On average, lower cell numbers were acquired for NPM1(1) samples than NPM1(2) and NPM1(3) samples (Figure S6a), and NPM1(3) samples had higher number of cells showing strong expression of multiple markers (Figure S6b).

Similar to single-cell RNA-seq data, we projected all 433,646 single-cells from the 16 *NPM1*-mutated AML samples acquired by spectral flow cytometry onto a UMAP plot and annotated the cells based on the *NPM1* subtype as measured by bulk transcriptomics. Like single-cell RNA-seq results, the data showed that cells from the three *NPM1*-mutated AML subtypes were differently distributed over the UMAP plot (Figure 3b), and that single-cells from the same *NPM1* subtype co-localized (Figure S6c). To further characterize the phenotype of the AML cells, we performed clustering with PhenoGraph,^20^ revealing 33 different cell clusters (Figure 3c, see Methods). Cell cluster abundances differed between the three NPM1 subtypes (Figure S6c-d). Various cell clusters were significantly more abundant in NPM1(1) samples (clusters 4, 7, 11, 14), whereas other cell clusters were mainly present in NPM1(3) samples (clusters 2, 10, 12, 17, 21, 22, 23) (Figure S6e), illustrating that the AML sample composition differed between the three NPM1 subtypes.

### NPM1 subtypes display different cell surface markers

Based on what is known about marker expression during normal hematopoiesis from hematopoietic stem cells to more mature myeloid cell populations^21–23^, we grouped the 33 flow cytometry clusters into 5 major AML phenotypes, i.e. Hematopoietic Stem and Progenitor Cell-like (HSPC-like) (CD34^+^CD38^-^Lin^-^), Early Progenitor-like (CD34^-^ CD117^+^CD45RA^+^Lin^-^, CD34^-^CD117^+^CD45RA^-^Lin^-^, CD34^-^CD117^-^CD45RA^+^Lin^-^), Late Progenitor-like (CD34^-^CD117^-^CD45RA^-^Lin^-^), Mono-like (CD14^+^CD11c^+^, CD16^+^CD11c^+^, CD68^+^CD11c^+^), and DC-like (HLA-DQ^+^CD11c^+^) AML cells. The Mono- and DC-like clusters were defined by expression of at least one lineage (Lin) marker, including CD14, CD16, CD68 and/or CD11c, whereas all progenitor cell clusters were negative for these lineage markers (Figure 3d).

To corroborate these cluster annotations and as a data quality check, we selected CD33^+^ or CD34^+^ cells from our single-cell transcriptomics data and associated the flow cytometry markers with their corresponding gene expression (Figure S6f, see Methods). All markers used to annotate the 5 major AML phenotypes showed significant correlations between gene and protein expression (Figure 3e, S6f). Having ensured that marker expression correlated with their corresponding genes, we next correlated the markers of the 33 flow cytometry clusters with the clusters found in scRNA-seq data (Figure S6g). This analysis yielded similar patterns with the same 5 major AML cell subsets in Figure 3d. Of note, high-mito clusters from single-cell RNA-seq data correlated with late progenitor-like cells (CD33^+^CD34^-^CD117^-^CD45RA^-^Lin^-^). Next, we projected the 5 major cell subsets onto the UMAP plot of the flow cytometry data (Figure 3f) and demonstrated their distribution within each of the 16 AML samples (Figure 3g).

The flow cytometry data supported single-cell RNA-seq results, demonstrating that AML cells with HSPC-like phenotype dominated in NPM1(2) samples, whereas NPM1(1) samples mainly contained AML cells with late progenitor-like phenotypes, and AML cells with more mature monocyte- or DC-like phenotypes were abundantly present in NPM1(3) samples. The only discrepant sample was AML11, which has been identified as NPM1(2) sample by bulk transcriptomics. Further inspection of this sample confirmed lower *KIT* (CD117) gene expression in this sample as compared to other NPM1(2) samples (Figure S6h), which may explain its resemblance to NPM1(1) by spectral flow cytometry. Overall, the spectral flow cytometry data confirmed that there is intra-patient heterogeneity in *NPM1*-mutated AML samples and that inter-patient heterogeneity largely overlapped with our identified transcriptional *NPM1* subtypes.

### HSC in *NPM1*-mutated AML can be malignant, but also healthy or pre-leukemic

Two cell clusters in the flow cytometry data resemble HSCs characterized by CD34^+^CD38^-^CD45RA^-^ (cluster 24) or CD34^+^CD38^--^CD45RA^+^ (cluster 27) expression (Figure 4a). Cluster 24 was mainly present in NPM1(1) samples (Wilcoxon rank-sum *P* = 0.048 and *P* = 0.024), whereas cluster 27 was more dominant in NPM1(2) samples, reaching the significance threshold when compared to NPM1(1) (Figure 4b, Wilcoxon rank-sum *P* = 0.05). To explore whether these cell clusters represent malignant or non-malignant cells, we made use of Cellular Indexing of Transcriptomes and Epitopes by Sequencing (CITE-seq) data from Sergi-Beneyto *et al.*^10^. We first selected the 7 *NPM1*-mutated AML samples from their cohort and re-integrated these samples. Next, we projected the cells on a UMAP plot colored by author’s original cell type annotations, which resulted in various clusters including lymphoid (B-, T- and NK-cells) and AML cells, which clearly clustered separately (Figure 4c). Besides lymphoid cell clusters, there was another healthy (or pre-leukemic) cell cluster identified (cluster 16), which consists of cells mostly lacking the *NPM1* mutation (Figure 4d, S7a). This cluster had high CD34 protein expression (Figure 4e, S7b), and was present in all 7 *NPM1*-mutated AML samples (Figure S7c). Since this cell cluster had the same phenotype as cell cluster 24 in our flow cytometry data (CD34^+^CD38^-^CD45RA^-^), our analyses suggest that cluster 24 represents healthy (or pre-leukemic) HSC that are mainly present in NPM1(1) samples.

**Figure 4.**
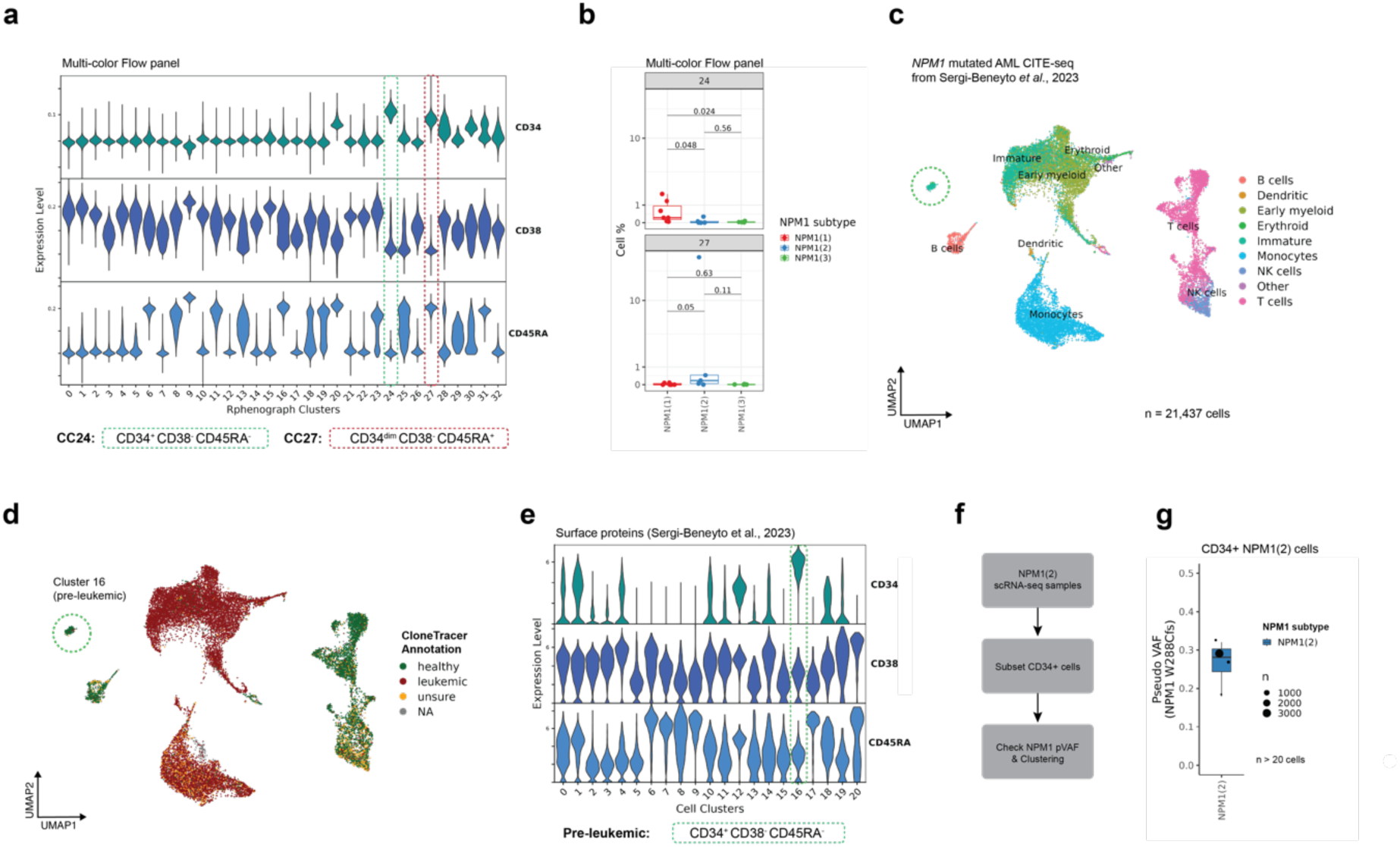
AML in NPM1(1) subtype contain healthy or pre-leukemic HSCs. **a** Cell surface expression of CD34, CD38 and CD45RA on all cell clusters as identified by spectral flow cytometry. Two CD34+CD38-cell clusters resembling HSC or early progenitor cells (CC24 and CC27) are highlighted. **b** Percentages of cells within each sample for CC24 and CC27 grouped by NPM1 subtypes. p-values were calculated using the Wilcoxon rank-sum test. **c** UMAP plot of integrated cells from 7 *NPM1-*mutated AML at diagnosis of Sergi-Beneyto *et al.*^1^ analyzed by CITE-seq. Cells colored by reported cell type annotations. **d** Same UMAP colored with CloneTracer annotations indicating malignant cells. Cluster 16 is highlighted as healthy or pre-leukemic cell subset. **e** Surface expression of CD34, CD38 and CD45RA as measured by CITE-Seq for all cell clusters. **f** Strategy for single-cell RNA-seq samples to show pVAF in CD34^+^ cells of NPM1(2) subtype. **g** NPM1^W288Cfs^ pVAF for CD34^+^ NPM1(2) cells.

To show that early progenies indeed carry the *NPM1* mutation in NPM1(2) samples, we selected CD34^+^ cells within this subtype (n=3,668) and demonstrated that the majority of these cells exhibit LT-HSC-like, LMPP-like or Progenitor-like phenotypes (Figure 4f, S7d), and that the mutant *NPM1* allele was detected across all different clusters and patients, thereby confirming that cell cluster 27 in the flow cytometry data (CD34^+^CD38^-^CD45RA^+^) represents malignant cells (Figure 4g, S7e).

In conclusion, these data support that AML cells in NPM1(2) samples closely resemble pluripotent progenitor cells, indicating that these cells are arrested early in hematopoiesis, whereas NPM1(1) samples are dominated by more committed progenitor cells, and NPM1(3) samples by AML cells with more mature monocyte- or DC-like phenotypes.

## DISCUSSION

*NPM1-*mutated AML accounts for ∼30% of adult AML patients and it is a single entity in recent WHO and ICC 2022 classification^1,2^. Nevertheless, previous studies demonstrated that this genetic subclass harbors significant phenotypic heterogeneity with potential prognostic and therapeutic significance^5,6,9^. Here, we followed up our recent work in which analyzed >1200 bulk transcriptomics data of AML cases and identified three major transcriptional subtypes of *NPM1*-mutated AML. In the current manuscript, we leveraged same-sample AML single-cell RNA sequencing and spectral flow cytometry data to show that these transcriptional subtypes can be traced back to differences in proportions of cell subsets, ranging from early hematopoietic stem or progenitor cells to more committed progenitors and mature myeloid cell types. Overall, the results suggest that in *NPM1*-mutated AML, myelopoiesis continues with malignant cells skewed towards different myeloid cell types.

Based on our single-cell analyses, we hypothesize that AML belonging to the NPM1(2) subtype originate from early HSC- or LMPP-like progenitor cells producing myeloid offspring. Flow cytometry data showed that stem cell marker CD34 as well as CD45RA and CD117, which are early progenitor markers in normal hematopoiesis^21^, are often present in leukemic cell populations in NPM1(2) subtype. scRNA-seq data also demonstrated that HSC-like and LMPP-like leukemic cells are most frequent in NPM1(2) subtype, and that all leukemic cell populations in this subtype have higher stem cell scores than leukemic cell populations in AML of the NPM1(1) and NPM1(3) subtypes. Since monocyte- and DC-like AML cells have also been detected in the NPM1(2) subtype, our data suggest that at least a proportion of the early progenitor-like AML cells retain the capacity to differentiate into more mature myeloid cells.

In contrast to AML belonging to NPM1(2), CD34, CD45RA and CD117 were absent or low on all or the majority of leukemic cell populations in NPM1(1) and NPM1(3) samples. Flow cytometry and scRNA-seq data showed that AML cells with more mature phenotypes, expressing high levels of maturation markers (CD11c, CD14, CD16, HLA-DR/DQ/DP, LILRB4, CD163, CD68), are most abundant in samples of the NPM1(3) subtype. This suggests that these AML may have developed from progenitor cells that are more primed to the monocytic lineage, such as GMP cells, monoblasts or promonocytes^21^, and that these progenitor cells, upon acquiring the *NPM1* mutation, retain the ability to differentiate into mature myeloid cells. An alternative possibility is that NPM1(3) samples originate from the same HSC- or LMPP-like cells as NPM1(2) samples, but the progenitor cells in NPM1(3) samples are less arrested and therefore produce more mature myeloid offspring. This possibility may be supported by our scRNA-seq data showing that besides mature monocyte-, macrophage- and DC-like AML cells, leukemic cells with HSC-, LMPP- and GMP-like phenotypes are also present in samples of the NPM1(3) subtype, albeit in smaller proportions than in NPM1(2) samples. Due to the abundant presence of mature AML cells, NPM1(3) cases could be more resistant to treatment with the BCL2-inhibitor Venetoclax, as a monocytic phenotype in AML has been associated with higher resistance to Venetoclax and lower overall survival rates ^9,24–27^.

Flow cytometry data also shows absence of CD34, CD45RA and CD117 on all or the majority of cell populations in AML of the NPM1(1) subtype, suggesting that these AML may have originated from cells that are more committed than HSC- and LMPP-like leukemic cells. Our scRNA-seq data shows that in particular cells with a high mitochondrial RNA content are abundantly present in the NPM1(1) subtype. A similar subset was recently reported by Naldini *et al.* for *NPM1-*mutated AML samples^13^. Our data shows evidence that AML cells with a high content of mitochondrial RNAs resemble progenitor cells that are more committed than HSC and LMPP, such as CMP, GMP or myeloblasts, yet lack the characteristic markers to define the cell type of origin.

Interestingly, the functionally complementary IDH1/2 and TET2 co-mutations, that are enriched in NPM1(1) subtype, have previously been linked to a disturbed alpha-ketoglutarate metabolism which leads to genome-wide hypermethylation^28^. This might explain upregulation of Fat mass and obesity-associated protein (FTO) gene expression as previously reported for the NPM1(1) subtype^5^, encoding a mRNA demethylase potentially counteracting this global hypermethylation. This suggests that cells with a high content of mitochondrial RNAs, which are notably abundant in NPM1(1) samples, may exhibit reduced expression of various cell surface markers as a result of extensive hypermethylation.

As described above, AML cells with a high content of mitochondrial RNA may represent late progenitor cells such as CMP- or GMP-like cells or myeloblast-like cells that are more committed to the granulocytic lineage^21^. The hypothesis that AML belonging to the NPM1(1) subtype may have originated from more differentiated progenitor cells than AML of the NPM1(2) subtype is supported by flow cytometry data showing a small healthy or pre-leukemic cell cluster similar in phenotype to healthy HSC (CD34^+^CD38^-^CD45RA^-^) in AML of the NPM1(1) subtype, whereas another small cell cluster resembling leukemic stem cells (CD34^+^CD38^-^CD45RA^+^) is found in AML of the NPM1(2) subtype. Kersten *et al.*^29^ also demonstrated that rare CD34^+^CD38^-^ cell populations in CD34-negative AML, which are often *NPM1*-mutated AML, are healthy or pre-leukemic HSCs, lacking expression of CD45RA and patient-specific mutations. However, the authors did not report on the presence of rare CD34^+^CD38^-^ leukemic stem cell populations expressing CD45RA that are present in AML in which the large malignant cell populations are negative for CD34. Our data showed that small healthy and leukemic CD34^+^CD38^-^ cell populations can both be present in *NPM1*-mutated AML, but that leukemic stem cells are more dominant in NPM1(2) samples, and healthy hematopoietic stem cells in NPM1(1) samples. Whether both cell populations are absent from NPM1(3) samples is unknown, since our small cohort of 16 samples included only 4 NPM1(3) samples. Therefore, deep analysis of a larger panel of *NPM1*-mutated AML is necessary to confirm our observations and draw definite conclusions.

The current study also has some limitations. First, in our previous analysis with bulk-transcriptomics, the size of the compendium allowed us to subdivide NPM1(2) and NPM1(3) each in two further subtypes^5^. For instance, NPM1(3) could be distinguished into two clusters dominated by CD14^+^ or CD16^+^ monocyte-like AML cells, indicating that there is still underlying heterogeneity, and that future single-cell studies are warranted to deduce the mechanisms underlying these additional NPM1 subtypes. Second, for variant calling, we recommend interpreting results from read-level pseudo VAF (pVAF) values on cell clusters, since variant calling on single-cells relies on only a few reads and may therefore be heavily biased and lead to wrong interpretations. Yet, pVAF values in our experiments were not affected by variable *NPM1* gene expression between different cell types. Furthermore, although we convincingly demonstrated that AML cells with a high content of mitochondrial RNAs have similar *NPM1* pVAF levels as other leukemic cell clusters, thereby confirming their malignant origin, our antibody panel used for spectral flow cytometry lacked any strong positive lineage markers for these cells. Our single-cell transcriptomics data provide an opportunity to refine our flow cytometry panel, and since the high mito clusters lack most cell surface markers, other flow cytometry protocols may be considered requiring cell fixation and permeabilization to allow inclusion of antibodies against RUNX1, FOXP1, PBX3, and MEIS1, which have been identified as transcription factors that are highly expressed. (see Supplementary Table 2).

In conclusion, our data showed that AML in the three transcriptional NPM1 subtypes previously identified by bulk transcriptomics contained different proportions of the same leukemic cell types resembling early progenitor or more mature myeloid cells. Our data favor a model in which AML of the three NPM1 subtypes originate from different progenitor cells with variable capacity to produce more differentiated offspring. This model suggests that *NPM1*-mutated AML belonging to different transcriptional subtypes may differ in susceptibility to standard or new anti-cancer therapies, and our model may thus have potential relevance for prognosis and treatment of patients with *NPM1*-mutated AML.

## METHODS

### AML samples

All sixteen peripheral blood and bone marrow samples were obtained from patients with AML at diagnosis (n=15) or relapse after chemotherapy (n=1) with written informed consent according to the Declaration of Helsinki. Mononuclear cells were isolated by Ficoll-Isopaque density gradient centrifugation and cryopreserved in the Leiden University Medical Center (LUMC) Biobank for Hematological Diseases, and approval to use *NPM1*-mutated AML for this research has been obtained by the LUMC Institutional Review Board (protocol no. RP24.056).

### Bulk RNA-seq data

The HAMLET^30^ pipeline was run on bulk RNA-sequencing data to screen mutations in 13 genes. Results for *NPM1* mutations and *FLT3-ITD*s have been validated by PCR on genomic DNA, followed by electrophoretic fragment size analysis. All subtype annotations for BEAT-AML and LUMC cohort with corresponding count data and meta tables (including genetic aberrations) can be found: https://osf.io/wq7gx/. In our previous work, we reported 5 NPM1 subtypes, here we re-merged these clusters into 3 major ones based on the clustering tree. We refer to Supplemental Figure 2 in Severens *et. al*.^5^ for more detailed view of different AML transcriptional subtypes.

### Sample preparation for single-cell RNA-seq

Single-cell RNA-seq was performed using the 10X Genomics Chromium Next GEM Single-cell 3′ Kit v3.1 (PN-1000268) and 10X Genomics Chromium Next GEM Chip G Single-cell Kit (PN-1000120) according to the manufacturer’s instructions (User Guide CG000315 Revision D). The libraries were pooled and sequenced on a NovaSeq 6000 platform (Illumina) using a 300-cycle kit S4 flow cell with v1.5 chemistry.

### Downstream analysis of single-cell RNA-seq data

*CellRanger* v7.0.0 was run on all samples with the human reference genome hg38. For all QC *Seurat* v4 was used^15^. Our QC pipeline had three steps per sample: 1) soft filtering, 2) low quality cluster removal, and 3) doublet detection. In soft filtering, *Seurat* objects were created with cells expressing at least 200 genes and with the genes expressed at least in 3 cells. Then, standard Seurat command list with default parameters was run to detect low quality clusters. Clusters with >15% mitochondrial and <1000 mRNA in median were removed. Next, *DoubletFinder* v3^31^ was run for each sample with 7.5% detection rate and then singlets were selected. To integrate the samples, CCA mode of *Seurat* pipeline was used with most variable 2000 features. Finally, after integration we filtered cells with >15% mitochondrial mRNA.

We used standard *Seurat* commands to scale and normalize the data on integrated features. The first 30 principal components were used to create UMAP plots. We used *clustree*^32^ to determine the optimal cluster number, based on *FindClusters* with resolutions sweeping from 0 to 1.2. We chose res=0.5, as clusters became stable (Figure S1a). Next, we merged two clusters (CC5 and CC12) into one GMP-like cluster as one of these clusters (CC12) had high expression of HSP-genes yet still retained its cell-type specific properties (Figure S1c, d).

### Annotation of single-cell transcriptomics clusters

We ran *Azimuth* to be able to project detailed cell type annotations from a healthy BM atlas. Most myeloid cells had low prediction scores whereas B- and T/NK-cells had higher scores. Therefore, we used annotations with >90% prediction score to faithfully annotate-like cell clusters. We also downloaded AML phenotype defining genes from van Galen *et al* ^12^. We performed differential expression analysis with MAST^33^ after removing ribosomal and HSP-related genes via one-vs-rest setting. Up-regulated genes were identified based on |logFC| > 0.5 and adjusted p < 0.05, and we further filtered out the genes from the list if they were expressed in less than half of the cells in the cluster of interest.

As two clusters – ‘High-mito’ & ‘High-mito (G2/S)’ - had relatively high mitochondrial content, we tried regressing out mitochondrial percentage with the *ScaleData* function to make sure that these clusters are not technical artifacts. To check whether these subsets might be attributed to integration artifacts, we re-run the standard *Seurat* pipeline per sample with default parameters on their raw RNA data and annotated all cells with their cell type annotations based on the integrated map, yet high-mito cells created their own cluster within all samples.

### Variant calling from single-cell-RNA-seq data

For hotspot mutations shown in Figure 1b (Supplementary Table 1), we first added safety margins 5bp to each side not to miss variants due to alignment issues. Then, *freebayes*^34^ was run on .bam files of each sample with >5 read support with human reference genome (hg38) to create .vcf files. Next, *pysam*^35^ was used to detect mutated- and wild type-reads from these called variants. Then, these reads were counted per cell barcode. The *AddMetaData* function was used to merge these cell barcodes with the integrated *Seurat* object.

### Analysis of CITE-seq data from literature

We downloaded the *Seurat* object of Sergi-Beneyto *et al.* through the figshare link in their manuscript: https://figshare.com/articles/dataset/Seurat_CloneTracer_Cohort/20291628. We selected the 7 *NPM1-mutated* samples from the cohort A patients as these were primary AML (A5, A8-A13). Next, we re-integrated these samples with their raw RNA assays using the *Seurat* CCA pipeline based on the most variable 2000 genes. The pre-leukemic subset (cluster 6 in the original manuscript) clustered separately from all other leukemic myeloid cells in our UMAP plot. Clustering was performed on the first 30 principal components with default parameters of *Seurat* pipeline.

### Stemness score calculations

We first created pseudobulk expression for all genes with the *AggregateExpression* from *Seurat* package per sample across cell clusters. Then, per pseudobulk we adjusted the library sizes of RNA counts with *cpm* function from *edgeR*^36^, and then log-transformed the counts. Next, we used stemness score genes and coefficients to calculate stemness scores of each pseudobulk. The stemness score of each pseudobulk is calculated based on its expression multiplied by their corresponding coefficients from Ng. *et al*.^19^.

### Spectral flow cytometry and down-stream analysis

AML patient derived bone marrow or peripheral blood mononuclear cells were thawed, purified by ficoll-paque and stained with a 23-color antibody panel on the same day as data acquisition. Cells were stained with Zombie Red viability dye (Biolegend) for 15 minutes at room temperature (RT), washed, incubated for 15 minutes in PBS supplemented with 2.5% human serum (Sanquin) and subsequently stained with fluorochrome-conjugated antibodies in Brilliant stain buffer plus (BD) for 30 minutes at 4°C. The following antibodies were included in the spectral flow cytometry marker panel: CD3-BV421 (UCHT1, BD), CD4-BV421 (OKT4, Biolegend), CD8-BV421 (SK1, Biolegend), CD11c-BV605 (3.9, Biolegend), CD14-V450 (M5E2, BD), CD16-BUV496 (3G8, BD), CD19-BV421 (HIB19, Biolegend), CD33-PE (WM53, Biolegend), CD34-APC (581, Biolegend), CD38-PE-Fire810 (S17015F, Biolegend), CD45-Spark Violet 538 (HI30, Biolegend), CD45RA-BV750 (HI100, Biolegend), CD61-BV650 (RUU-PL7F12, BD), CD68-PE-Vio615 (REA886, Miltenyi), CD117-PE-Cy5 (104D2, Biolegend), CD163-BUV661 (GHI/61, BD), GPR56-BUV563 (CG4.rMAb, BD), CLL-1-BUV805 (50C1, BD), LILRB4-BUV737 (ZM3.8, BD), pan-HLA class 1-PerCP (W6/32, Biolegend), HLA-DP-BV786 (B7/21, BD), HLA-DQ-FITC (HLADQ1, Biolegend), HLA-DR-APC-Fire810 (L243, Biolegend). Data were acquired using a 5-Laser Aurora (Cytek Biosciences) using SpectroFlo software (v2.0, Cytek). After acquisition, data were compensated, scaled by applying cofactors and gated on viable myeloid cells expressing CD33 and/or CD34. Lymphocytes were excluded based on expression of CD3, CD4, CD8 or CD19. Compensation, scaling and gating was performed using OMIQ software (Dotmatics), after which single-cell protein expression data for 433,646 cells was exported.

For the down-stream analysis, 19 differentiation markers were selected (CD117, CD11c, CD14, CD16, CD163, CD33, CD34, CD38, CD45, CD45RA, CD61, CD68, CD85k-LILRB4, CLL-1, GPR56, HLA class 1, HLA-DP, HLA-DQ, HLA-DR). The RPhenoGraph pipeline was run with reported parameters in the manuscript (k = 60), which created the final 33 flow-based cell clusters.

To project the single-cell RNA-based labels of cell clusters on the flow cytometry data, we first subsetted CD33 or CD34 expressing cells in the single-cell RNA-seq data and selected common markers between both modalities. Next, the median expression of each marker was scaled and centered among clusters to be comparable in both modalities. Next, we calculated the Pearson correlation coefficients across all cell clusters of flow cytometry and single-cell RNA-seq data.

## DATA AVAILABILITY

All single-cell RNA-sequencing data was submitted to EGA with the accession number EGAS50000000332, and they are accessible upon request (due to privacy rules). Processed version of the scRNA-seq data for all 16 *NPM1*-mutated AML samples were uploaded to Figshare under the DOI: https://doi.org/10.6084/m9.figshare.26189771.

## CODE AVAILABILITY

All source codes are available at https://github.com/eonurk/scNPM1. Our AML variant calling pipeline for single-cell RNA-seq data is also open-sourced and available under the same repository.

## Data Availability

All raw files for single-cell RNA-sequencing data was submitted to EGA with the accession number EGAS50000000332, and they are accessible upon request (due to privacy rules). Processed version of the scRNA-seq data for all 16 NPM1-mutated AML samples were uploaded to Figshare under the DOI: https://doi.org/10.6084/m9.figshare.26189771.

https://osf.io/wq7gx/

https://doi.org/10.6084/m9.figshare.26189771

## ACKNOWLEDGEMENTS

Authors would like to thank Peter van Balen for his help on data acquisition and transfer, and Leon Mei and Davy Cats for their help on data upload. This study received financial support from a strategic investment from the Leiden University Medical Center, integrated within the Leiden Oncology Center and conducted within the Leiden Center for Computational Oncology, and partly by the Dutch Cancer Society (project number 15152). EBA was supported by a personal grant from the Dutch Research Council (NWO; VENI: 09150161810095). The funding entities played no part in determining the study’s design, data collection, analysis, interpretation, manuscript composition, or the decision to submit it for publication. *BioRender* icons were used in graphical abstract, Figure 1a, 3a.

## AUTHOR CONTRIBUTIONS

EBA, MG, MJTR designed the project; EBA acquired funding; EMA, NES, MG, SK and HV helped with data generation and transfer; EOK, EMA performed computational and statistical analyses; EOK created all figures; EOK, RSHZ implemented variant calling pipeline; EOK, EMA, RSHZ, JFS, EBA, MG, and MJTR supported data exploration and interpretation; MJTR, MG and EBA provided supervision and scientific direction; EOK, EBA and MG wrote the manuscript; and all authors critically reviewed the manuscript and figures.

---

**Figure S1.**
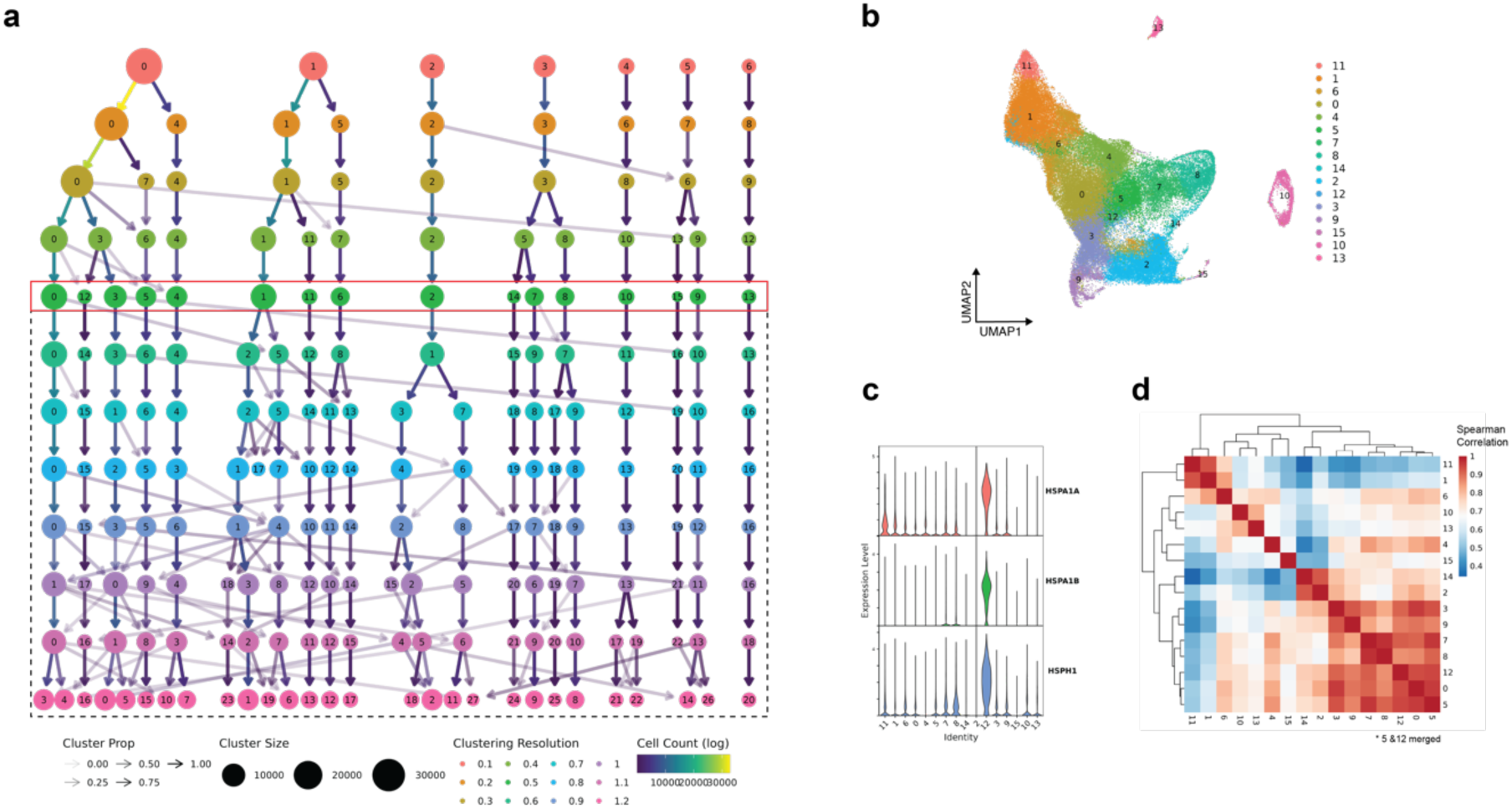
Distinct cell clusters in *NPM1*-mutated AML by single-cell RNA-sequencing. **a** Clustering tree for determining optimum clustering resolution. Selected resolution (r=0.5) is highlighted via red lines. **b** UMAP plot of *NPM1*-mutated AML atlas colored by clusters at selected resolution. **c** Heat shock protein (HSP) genes across cell clusters, indicating presence of a stress response probably due to technical artifacts in CC12. **d** Spearman correlations among cell clusters. Note that CC12 and CC5 were merged in further downstream analysis to recapitulate biology rather than technical artifacts, resulting in a total of 15 cell clusters.

**Figure S2.**
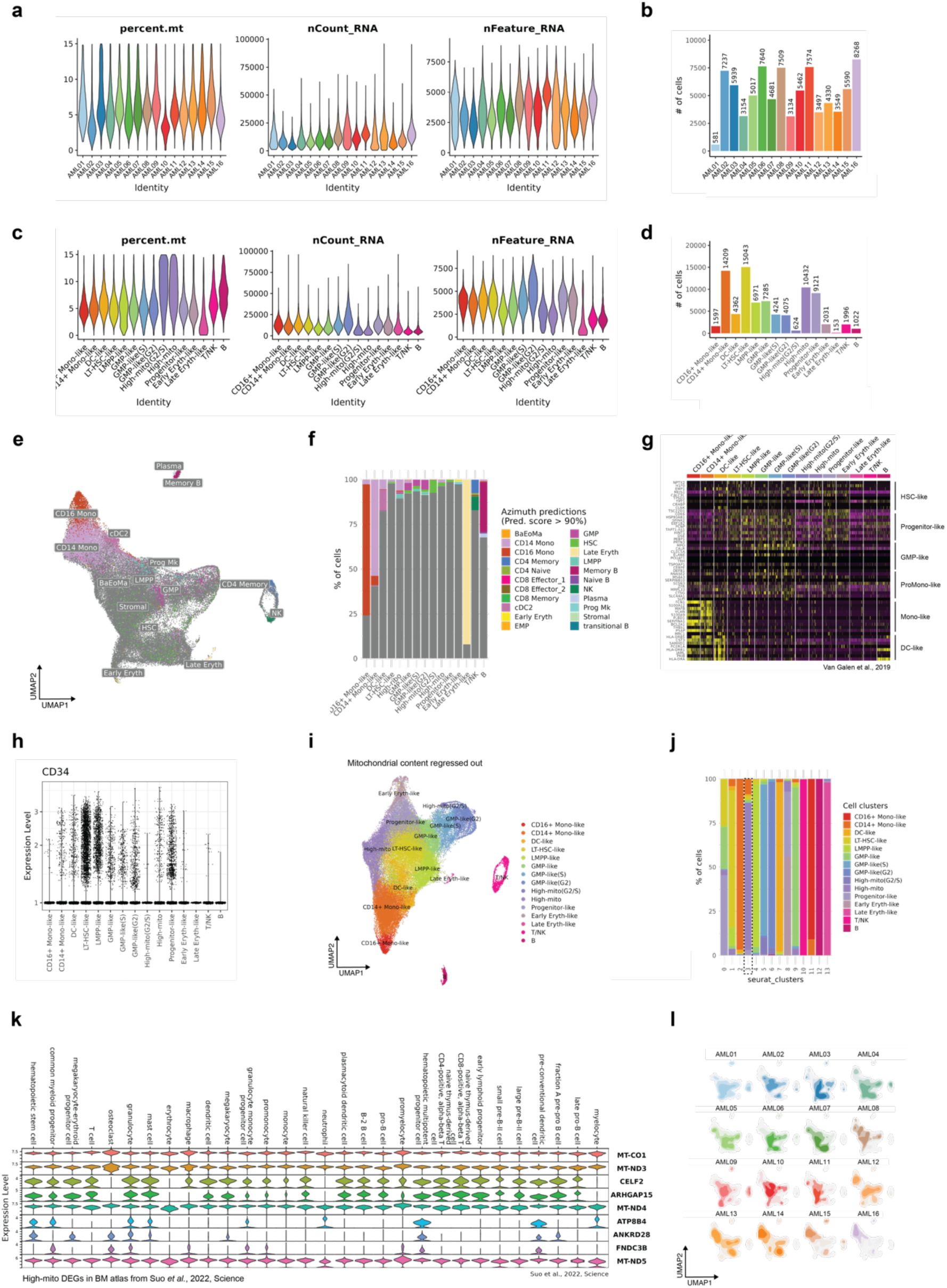
Quality control and annotation of cell clusters in *NPM1-*mutated AML by single-cell RNA-sequencing. **a, c** Standard quality control (QC) metrics for all AML samples and cell cluster. **b, d** Barplot for the number of cells per patient and cell cluster. **e** UMAP plot of *NPM1*-mutated AML atlas annotated by Azimuth at > 90% prediction score **f** Barplot showing the distribution of Azimuth annotations per cluster **g** Heatmap of AML cell type-specific genes as identified by van Galen *et al*.^2^ per cell cluster **h** *CD34* expression across cell clusters **i** UMAP plot of the *NPM1*-mutated AML cell atlas after regressing out high mitochondrial RNA content **j** Barplot showing the distribution of cell type annotations after correction for high mitochondrial RNA content. Cell clusters characterized by a high content of mitochondrial RNAs do not merge with other cell clusters after regressing out the high content of mitochondrial RNAs. **k** Expression of DEGs specific for cell clusters with a high content of mitochondrial RNAs in an independent cohort of healthy bone marrow samples of Suo *et al*.^3^ **l** Distribution of all cells for each separate AML sample on the *NPM1*-mutated AML cell atlas.

**Figure S3.**
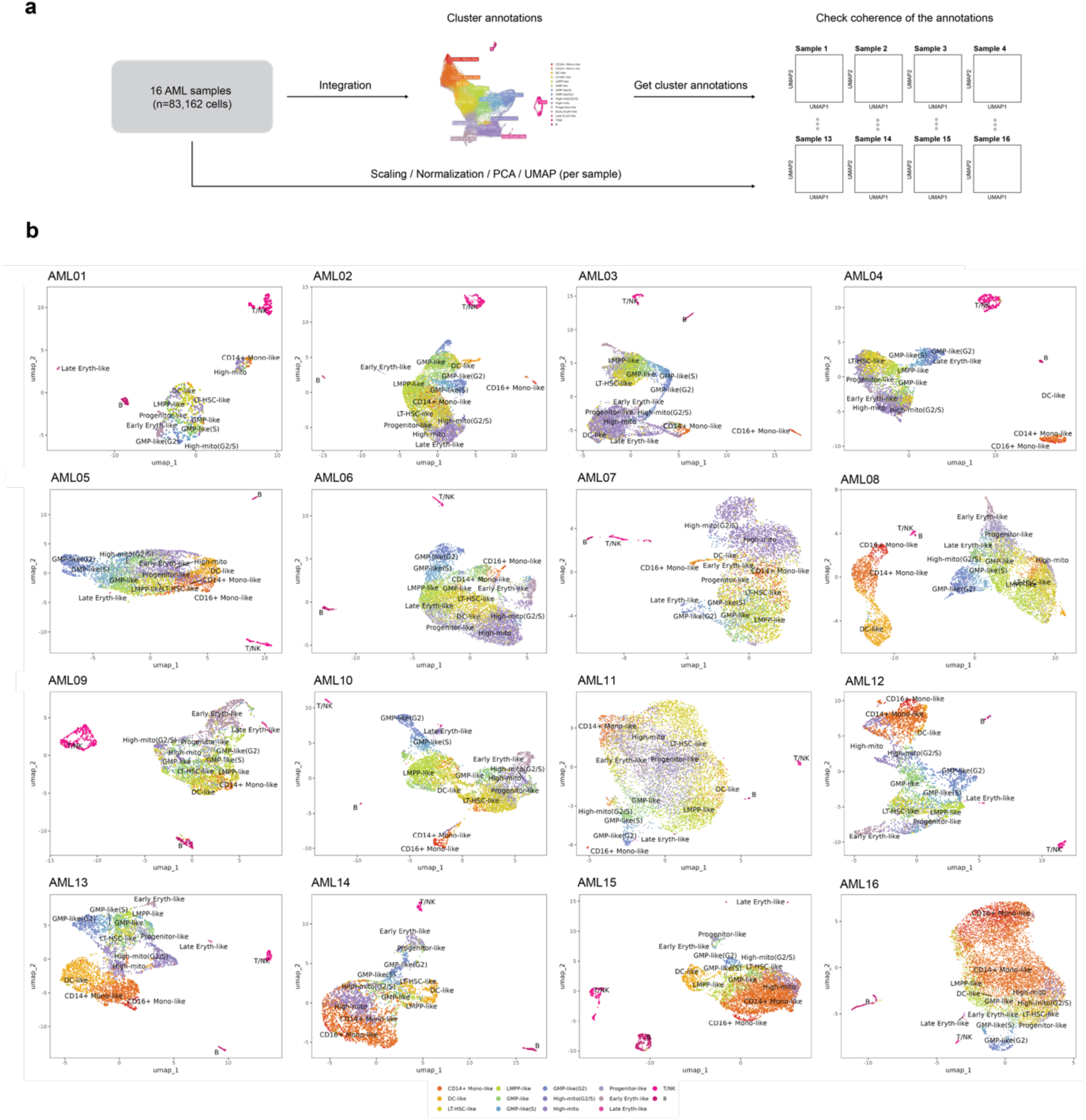
Quality control for integration and high-mito subsets. **a** Sketch showing the performed procedure for analyzing quality of integration. Seurat pipeline was run 16 AML samples independently. Then each sample’s cells were colored by the integrated cell type annotations to check for inconsistent cell clusters that merged randomly **b** UMAP plots showing all *NPM1*-mutated AML samples, cells colored by cell annotations from integrated cell atlas.

**Figure S4.**
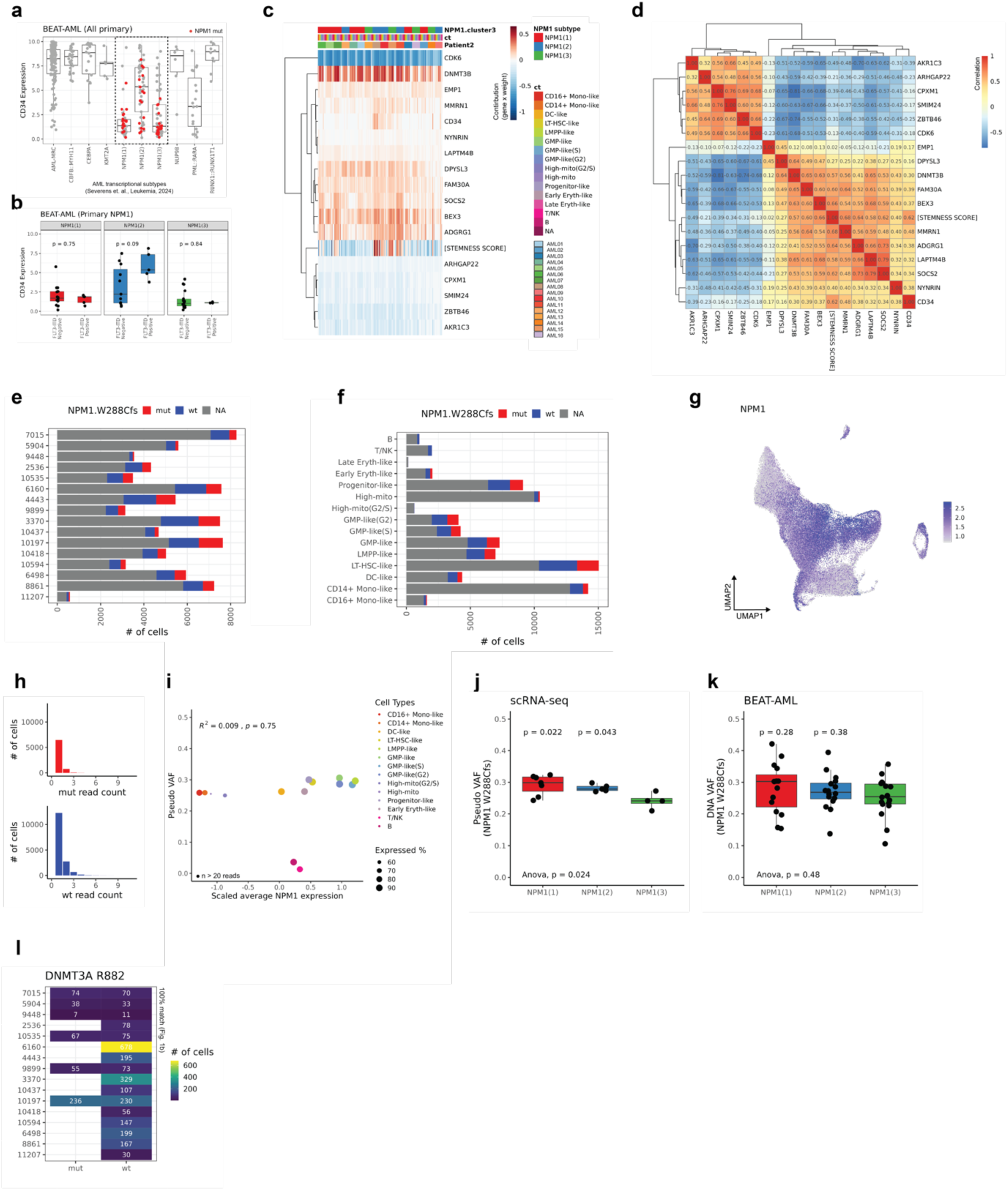
Detection of stemness and mutant alleles in NPM1 subtypes by single-cell RNA-sequencing. **a** *CD34* gene expression in AML samples at diagnosis of the BEAT-AML cohort. NPM1 subtypes are highlighted via a dashed line. Red dots indicate presence of the NPM1 mutation. **b** *CD34* gene expression across NPM1 subtypes in the BEAT-AML cohort divided based on FLT3-ITD status. **c** Contribution (gene expression x coefficient of that gene from Ng*. et al*^4^) of individual genes to the stemness score for each pseudo-bulk. 15 pseudo-bulks profiles were created for each sample (one per cell type) from single cell RNA-seq data. [Stemness Score] in the heatmap indicates the final stemness score for each pseudo-bulk. **d** Correlation of each gene with stemness score. Note that *CD34* expression is strongly correlated with stemness score (R = 0.62). *MMRN1* and *ADGRG1* also showed strong correlations with stemness score with R = 0.68 and R = 0.64, respectively. **e** Barplot showing the percentage of cells with mutated (red) or wild-type (blue) NPM1 alleles for each patient **f** and cell cluster. Gray indicates that no reads were found for NPM1. **g** *NPM1* expression on the UMAP plot. Note that *NPM1* expression is low for monocyte-like AML cells and AML cells with a high content of mitochondrial RNAs, and that this affects the sensitivity to call mutant NPM1 alleles in these cell clusters. **h** Distribution of mutant (mut) and wildtype (wt) reads in individual cells. Most cells have been annotated as mutant or wildtype based on one single read. **i** NPM1^W288Cfs^ pVAF with *NPM1* expression per cell cluster. **j** NPM1^W288Cfs^ pVAF across NPM1 subtypes **k** VAF_DNA_ for *NPM1*-mutated AML at diagnosis of the BEAT-AML cohort split by NPM1 subtype. p-values were calculated using the Wilcoxon rank-sum test. **l** Heatmap showing the number of cells with detectable reads for mutated or wild-type DNMT3A allele. Note that mutant DNMT3A alleles have only been detected in AML samples that are positive for the respective mutation as shown in Figure 1b.

**Figure S5.**
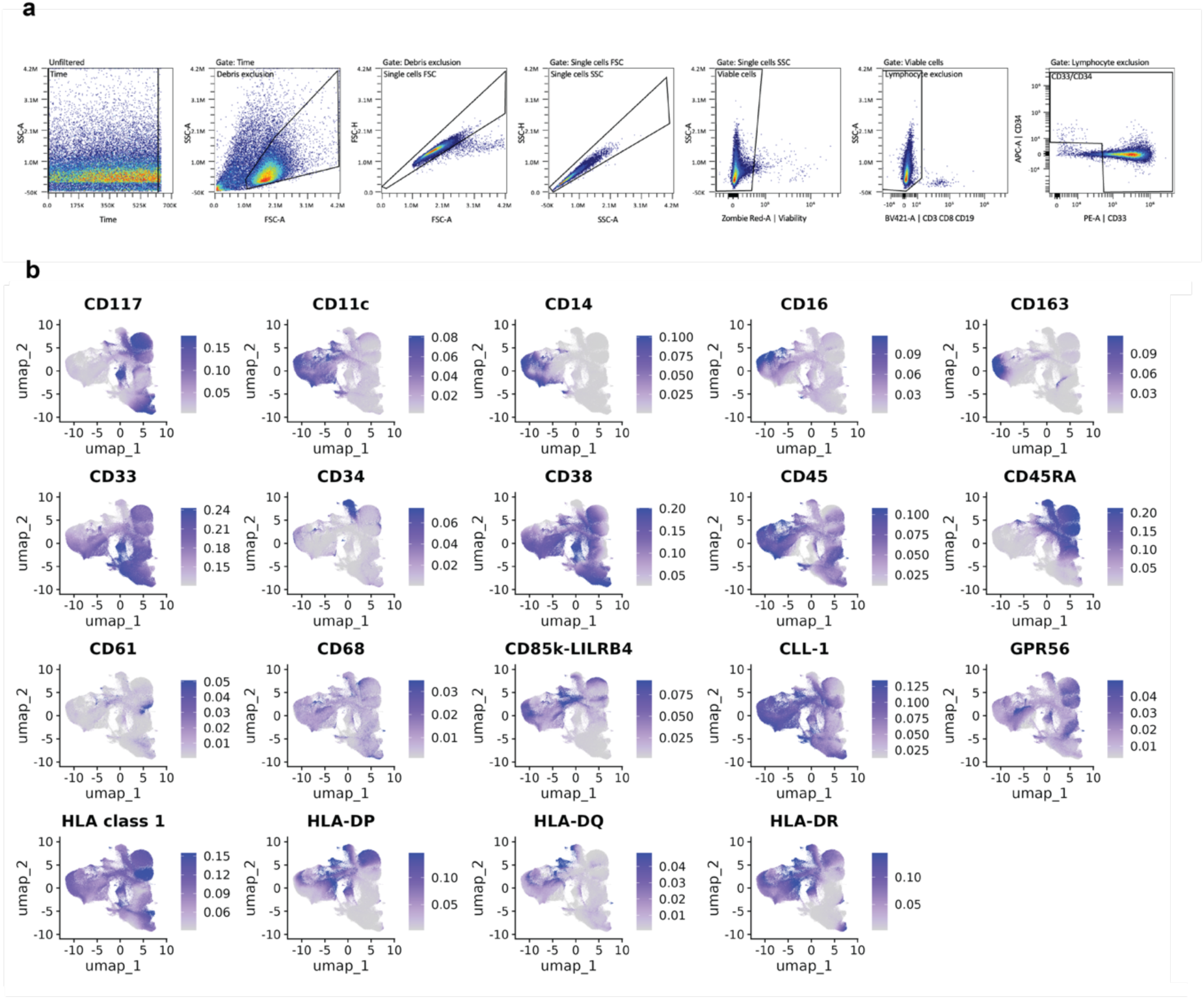
Marker expression in cell clusters by spectral flow cytometry. **a** Gating strategy for a representative AML in flow cytometry analysis. **b** UMAP plots showing expression of the 19 markers in cell clusters as measured by spectral flow cytometry. For each marker, maximum values were capped at 95% quantile.

**Figure S6.**
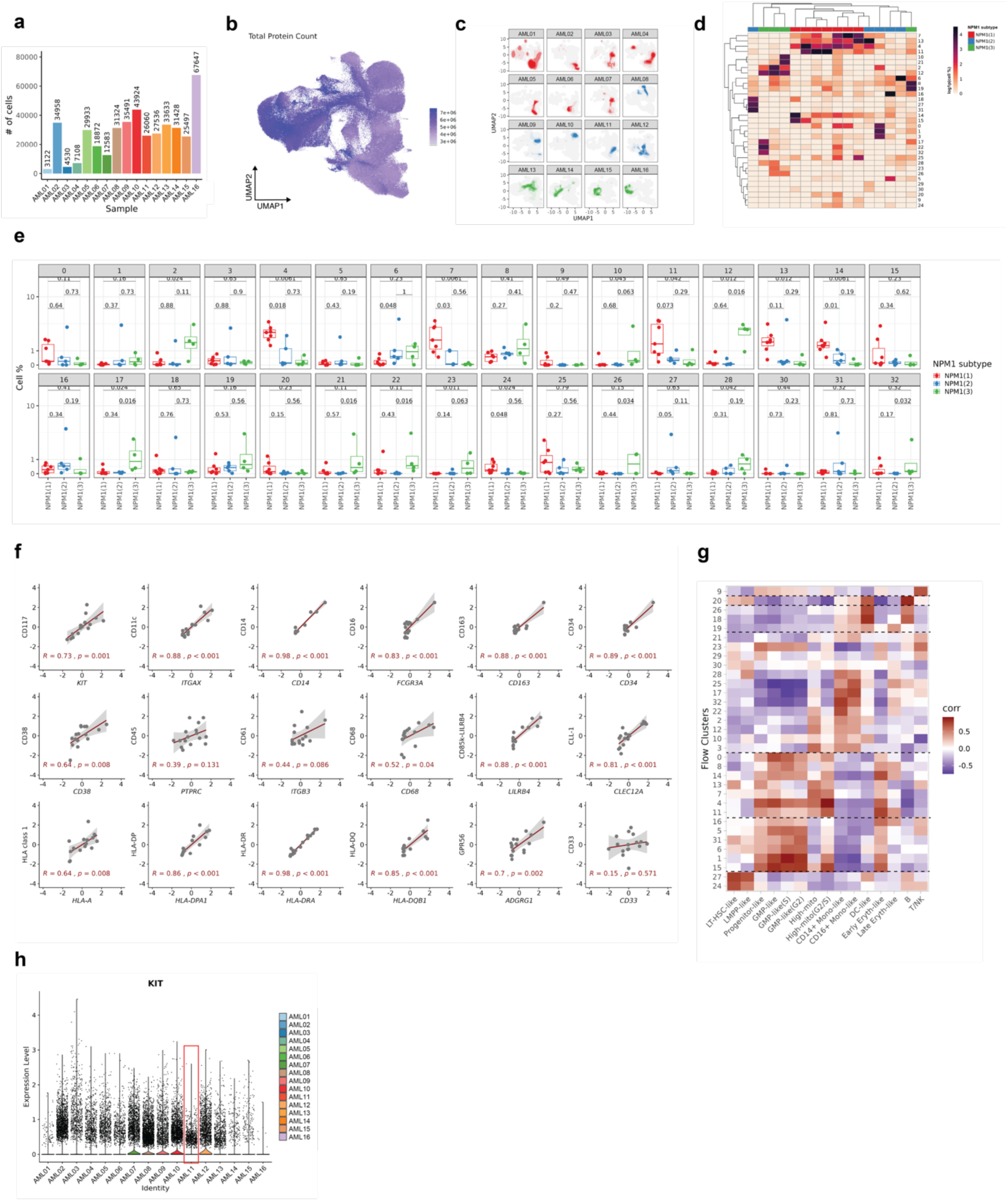
AML heterogeneity between NPM1 subtypes by spectral flow cytometry and single-cell RNA-sequencing. **a** Number of cells measured by spectral flow cytometry per AML sample **b** UMAP plot showing total expression of all 19 markers as measured by spectral flow cytometry **c** Distribution of cells on the UMAP plot for each AML sample using the *ggdensity* package. **d** Heatmap showing clustered percentages for each cell cluster per AML sample. Cell percentages were transformed with *log1p* function for better representation. **e** Proportion of individual cell clusters in samples across NPM1 subtypes. Pairwise p-values were calculated using the Wilcoxon rank-sum test. **f** Scatter plots showing normalized cell surface expression by spectral flow cytometry (y-axis) versus corresponding gene expression by single-cell RNA-sequencing (x-axis) for each sample. **g** Similar to (f) showing the proportion of individual cell clusters in samples by spectral flow cytometry versus single-cell RNA-sequencing on a heatmap plot. **h** *KIT* gene expression for each sample by scRNA-seq. AML11 has lower *KIT* gene expression than other NPM1(2) samples.

**Figure S7.**
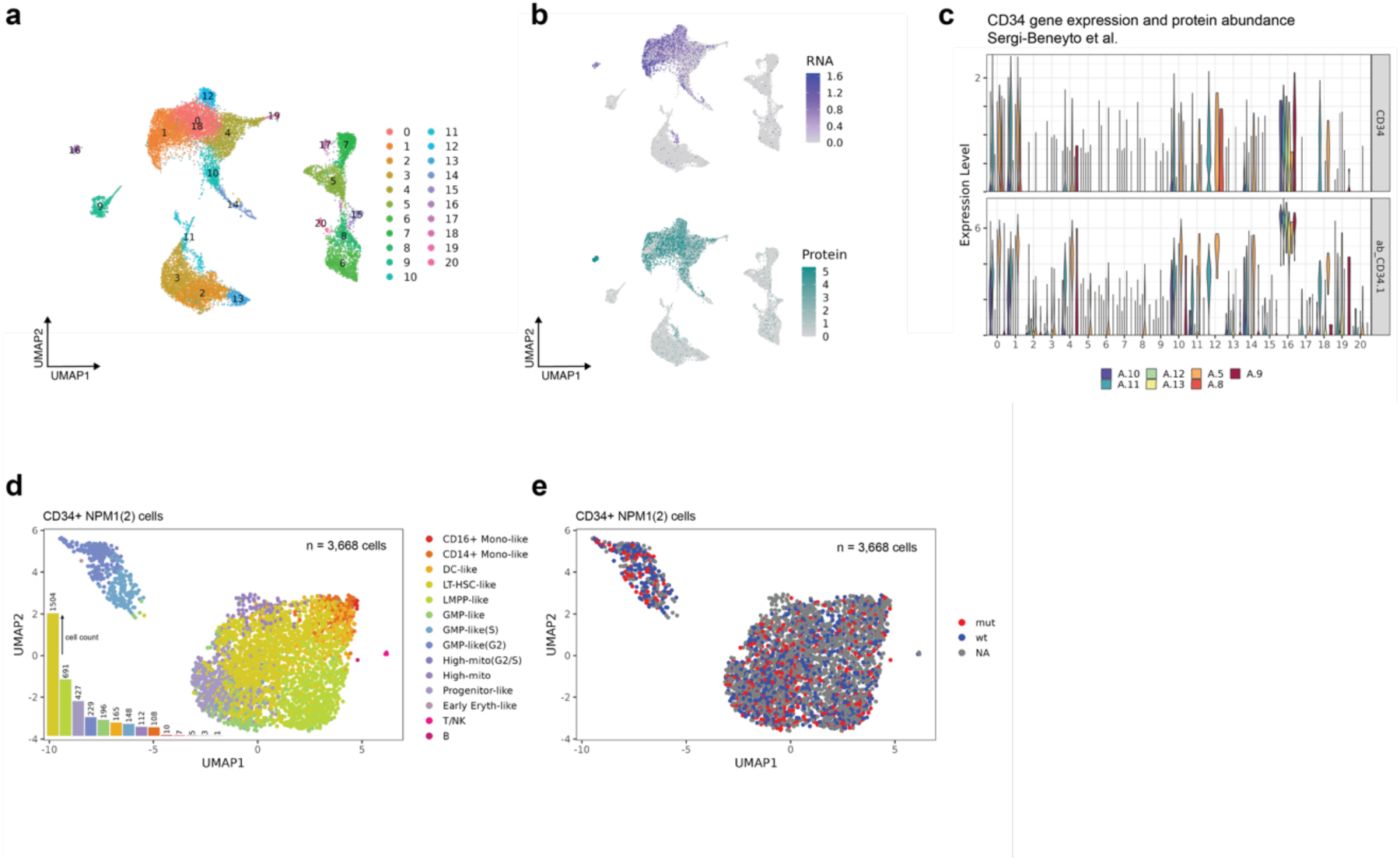
Analysis of an independent cohort of *NPM1* mutated AML. **a** UMAP plot of integrated cells from 7 *NPM1-*mutated AML at diagnosis of Sergi-Beneyto *et al*.^1^ analyzed by CITE-seq. Cell clusters are represented by different colors. **b** mRNA expression and surface expression of *CD34* by CITE-seq. **c** Similar to (b) with a violin plot and split per sample. Note that CD34 surface expression is consistently high for CC16 across samples. **d** *CD34*^+^ cells from NPM1(2) samples in our own AML cohort analyzed by scRNA-seq colored by each cell type. For each cell type, numbers of *CD34*^+^ cells are shown. *CD34*^+^ cells are most abundant in LT-HSC-like and LMPP-like AML cells. **e** The mutant NPM1^W288Cfs^ allele is present in all cell types identified in *CD34*^+^ NPM1(2) cells.

